# A retrospective cross-sectional analysis of community perceptions of flu and COVID-19 vaccines at Turtle Creek Primary Care Center

**DOI:** 10.1101/2022.04.15.22273479

**Authors:** Anjana Murali, Jorna Sojati, Marina Levochkina, Catherine Pressimone, Kobi Griffith, Erica Fan, Allie Dakroub

## Abstract

**Background:** Influenza (flu) and COVID-19 vaccination rates are subpar across the US, especially in racial and/or socioeconomic minority groups who are understudied in public health literature.

**Objective:** The objective of this study was to elucidate the attitudes of Turtle Creek patients towards flu and COVID-19 vaccines, with the goal of establishing targetable vaccine education gaps and ultimately increasing vaccine uptake in the community.

**Design/Patients:** This study was conducted as a retrospective cross-sectional analysis. Authors completed 123 patient phone surveys of patients cared for at the Turtle Creek Primary Care Center inquiring about flu and COVID-19 infection status and vaccination uptake (August 26 - October 10, 2021).

**Approach/Key Results:** Our data revealed a significant association between COVID-19 and flu vaccine acceptance. Additionally, we found a strong association between vaccine acceptance and age, with older patients being more likely to be vaccinated against COVID-19. Using multivariable logistic regression models, we assessed how flu and COVID-19 vaccine acceptance was affected by informational sources participants trusted most. In the COVID-19 models, those who cited “trusting medical professionals” had higher odds of vaccine acceptance while participants who cited “trusting social media” had significantly decreased odds of vaccine acceptance.

**Conclusion:** Our study revealed significant trends for flu and COVID-19 vaccine acceptance by sociodemographic factors and trust in the medical system. Using these data, we can create future interventions to overcome vaccine hesitancy.

## INTRODUCTION

Almost two years into the COVID-19 pandemic, consensus remains that the primary way to combat COVID-19 cases, hospitalizations, and deaths is to receive an approved vaccination. There are three current vaccines approved for use in the United States: Pfizer, Moderna and Johnson & Johnson, and at this time, all vaccine trials continue to expand age availability and combat virus variant changes. As boosters are introduced to increase immunity, the question remains: will the COVID-19 vaccine become an annual recommendation similar to the Influenza (flu) vaccination recommendation?

Early surveys and studies indicated that a large percentage of Americans were hesitant about COVID-19 vaccinations. A survey conducted in June of 2020, 6 months prior to any vaccine approval, determined that only 52% of Americans were “very likely” to receive a COVID-19 vaccination, while 15% indicated they were not likely and 7% responded they would definitely not receive a COVID-19 vaccination (1). Vaccine hesitancy is not unique to COVID-19. In a survey of nurses investigating flu vaccine uptake and plans to receive to COVID-19 vaccine, only 49% of respondents annually receive their flu vaccine and only 63% of respondents indicated their plans to receive a COVID-19 vaccine. This 63% is below the threshold for what is considered “herd immunity” (2). For the 2020-2021 flu season, there was an estimated 59.0% intent to receive the flu vaccine, with the highest intention found in those over the age of 65 (3).

Flu and COVID-19 vaccination rates are subpar across the US, and minority groups are even less likely to receive vaccinations compared to their counterparts (1,4–6). Only 41.2% of Black Americans and 38.3% of Hispanic Americans ages 18 years or older received a flu vaccine in the 2019-2020 season (6). During the COVID-19 pandemic and the 2020-2021 flu season, flu vaccination rates in Black and Hispanic populations remained stagnant (6), leaving nearly 60% of Black and Hispanic Americans unprotected and vulnerable to contracting the flu. COVID-19 vaccination rates among racial minorities remain unsatisfactory as well. Only 51% of Black Americans and 56% of Hispanic Americans received one COVID-19 vaccination dose as of December 2021, and it is likely that even fewer are fully vaccinated or boosted, meaning that nearly 50% of Black and Hispanic populations are exposed to significant morbidity and mortality risk associated with COVID-19 infection (5). These substandard vaccination rates emphasize the importance of understanding vaccine decision making in underserved and minority communities.

Many communities in Western Pennsylvania are primarily composed of racial and socioeconomic minority groups that may be experiencing deficiencies in COVID-19 and flu vaccination rates. The community of Turtle Creek borough, a suburb to the east of Pittsburgh, is a prime example: 34.6% of the people living in Turtle Creek identify as African American or biracial per the 2020 US census (7), and 33.6% of the Black population in Turtle Creek live below the poverty line (7). The University of Pittsburgh Medical Center (UPMC) operates the Turtle Creek Primary Care Clinic, a general medicine and pediatrics office in Turtle Creek that has a population of 71% Black and 6% Hispanic patients (7). For comparison, the population of nearby Pittsburgh is only 26% Black and 3.2% Hispanic (8). Turtle Creek’s community demographics plus the authors’ anecdotal experiences at the Turtle Creek clinic have raised questions about whether the patients at the Turtle Creek Clinic are hesitant to receive flu and COVID-19 vaccines. This study was created to elucidate Turtle Creek patients’ attitudes regarding COVID-19 and flu vaccines and personal vaccine hesitancy, with the goal of more effectively meeting the community’s vaccine needs.

## METHODS

### 1. Study Design and Cohort Description

The study design and survey were given approval by the Quality Improvement Review Committee at the University of Pittsburgh Medical Center (UPMC). The UPMC Quality Improvement Committee reviewed this project and deemed that it did not meet the federal definition of research according to 45 CFR 46.102(l) and did not require additional Institutional Review Board (IRB) oversight. The ethics of this study were also reviewed by the UPMC Quality Improvement Committee and deemed exempt from IRB review.

The study was conducted as a retrospective cross-sectional analysis. A random number generator was used to identify the 500 patients from the Turtle Creek Primary Care Clinic’s patient database. Contact information for these patients was curated by the Turtle Creek Clinic Manager. Using the Doximity Dialer displaying the clinic’s home phone number, authors made up to three attempts to contact each patient by their home phone number.

Authors were able to contact 176 participants, of which 123 verbally consented to the study and completed the survey. Surveys were primarily administered and collected between August 26th and October 10^th^, 2021, with 7 surveys completed April 17-May 23^rd^ during the study trial period.

The full survey questionnaire and answer options can be found in the **appendix (A1)**.

### 2. Statistical Analysis

#### 2.1 Dichotomization and Bivariate Associations Between Demographic Variables and Vaccine Acceptance

Statistical analyses were conducted in R, version 4.1.1, Vienna, Austria (9) and RStudio (10).

Descriptive statistics—mean, median, interquartile range (IQR)—were used for ordinal and continuous variables while measures of frequency were used for categorical variables. Non-parametric Mann-Whitney or Kruskal Wallis U tests were conducted for continuous and ordinal variables, while chi-square tests for independence and Fisher’s Exact tests were used for comparing categorical variables where appropriate. All tests were two-tailed when relevant with an α = 0.05.

#### 2.2 Discrete Binning of Demographic and Outcome Variables

For ease of statistical interpretability, age, gender, race, and education were binned. Age was binned in four brackets: ages (in years) 1) 18-24, 2) 25-44, 3) 45-64, and 4) >65. Gender was dichotomized as 1) Female, and 2) Not Female. Race was binned into three categories: 1) Black or African American 2) White, and 3) Non-Black or White. Finally, education was trichotomized into 1) some high school to high school diploma or GED, 2) Associates degree to some college, and 3) Bachelor’s degree completion or above.

COVID-19 vaccine acceptance status was dichotomized such that *vaccine acceptance* included participants who either answered that they 1) received the COVID-19 vaccine or 2) have plans to receive it while the *vaccine-hesitant* group consisted of participants who answered 1) no or 2) still thinking about it. History of flu vaccine acceptance was also dichotomized wherein *consistent flu vaccine acceptance* were participants who answered 1) always or 2) most years while the *flu vaccine-hesitant* group were participants who answered 1) few years or 2) never.

#### 2.3 Assessing Trusted Informational Sources on Vaccine Acceptance: Multivariable Regression Models

In order to assess the impact of types trusted informational sources on COVID-19 vaccine acceptance, two multiple logistic regressions models were run. Independent variables included trust in 1) *paper or digital news*, 2) *medical professionals/health organizations*, 3) *social media*, and 4) *word of mouth* which were all added to the model *a priori* (**A1**). The outcome of interest was the aforementioned dichotomized COVID-19 vaccine acceptance variable.

An identical methodology was performed for two additional models, now assessing trusted informational sources for flu vaccine acceptance status. Odds Ratios (OR) and 95% Confidence Intervals (CI) were reported in tabled results to indicate effect size for all four models.

### 4. Data Availability

The datasets analyzed during the current study are available from the corresponding author on reasonable request.

## RESULTS

Of the 2,556 patients at the Turtle Creek Primary Care Center in Turtle Creek, PA, we attempted to reach 400 patients for this study during the period of August 26th - October 10th, 2021. Seven of the surveys were administered during the trial period of April 17 - May 23, 2021, but since that data did not significantly differ from the rest of the surveys, they were included with the total study cohort. During the study period, COVID-19 vaccination guidelines stated that all adults, including pregnant women, were eligible for the vaccine and were recommended to be vaccinated (starting April 19, 2021). Of the 400 patients who were called, 224 patients were unreachable to our calls and 53 patients declined to participate, giving us a final study cohort of 123 patients (5% of the Turtle Creek patient population), as seen in **Figure 1**. In consenting patients for the study, we informed them that the study was done on a voluntary basis and their participation to our questions are not reported to their physicians.

**Figure 1.**
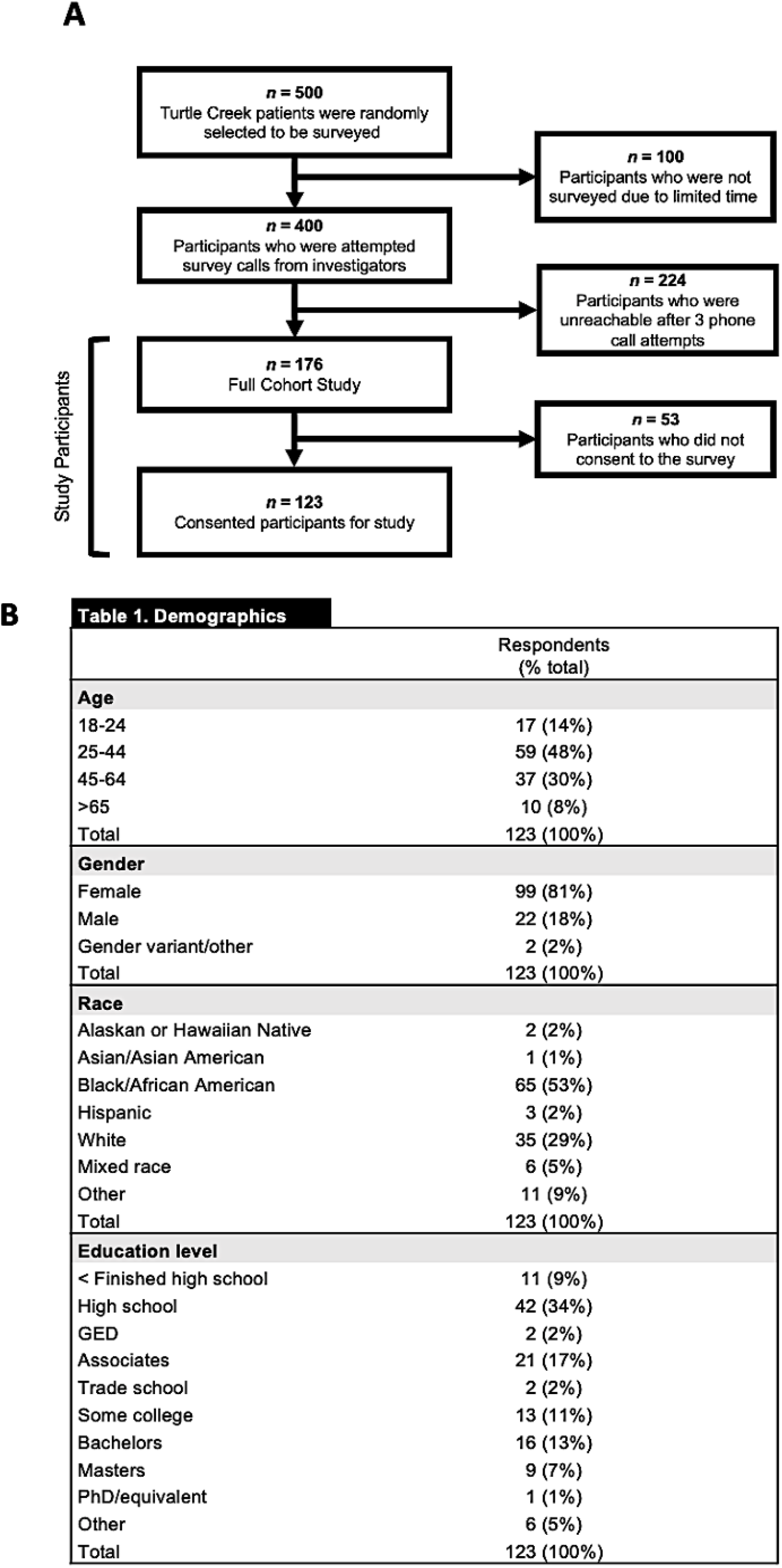
A) Flow chart representation of study design. Of 500 randomly selected patients of the Turtle Creek Primary Care Center, 129 participants were consented and are represented in this study. B) Table (Table 1) representation showing demographic information of the 129 consented participants.

Details of our study demographic can be found in **Table 1, Figure 1**. Of note, a majority of the survey participants identified as Black or African American (53%). Additionally, most of the participants identified as female (81%). 71% of the Turtle Creek Primary Care Clinic patient roster identifies as Black or African American.

At the point of surveying, the majority of participants had not been infected with the flu (98.3%) during the 2020-21 season (**Figure 2A)**. 10.9% of participants had received a COVID-19 diagnosis at this point (**Figure 2B)**. When looking at flu vaccination at large amongst this study cohort, one-third of patients reported routinely never getting the flu vaccine (**Figure 2C)**. In fact, less than one-third of the total cohort reported consistent flu vaccine uptake. Of note, flu vaccination status for 2021 was not assessed as it was not yet flu season. Meanwhile, COVID-19 vaccination uptake was much higher, with 53% of participants having already gotten the vaccine and another 6.1% having plans to get their shot (**Figure 2D)**. However, about one-third of the population did not want or did not have plans to receive the COVID-19 vaccine.

**Figure 2.**
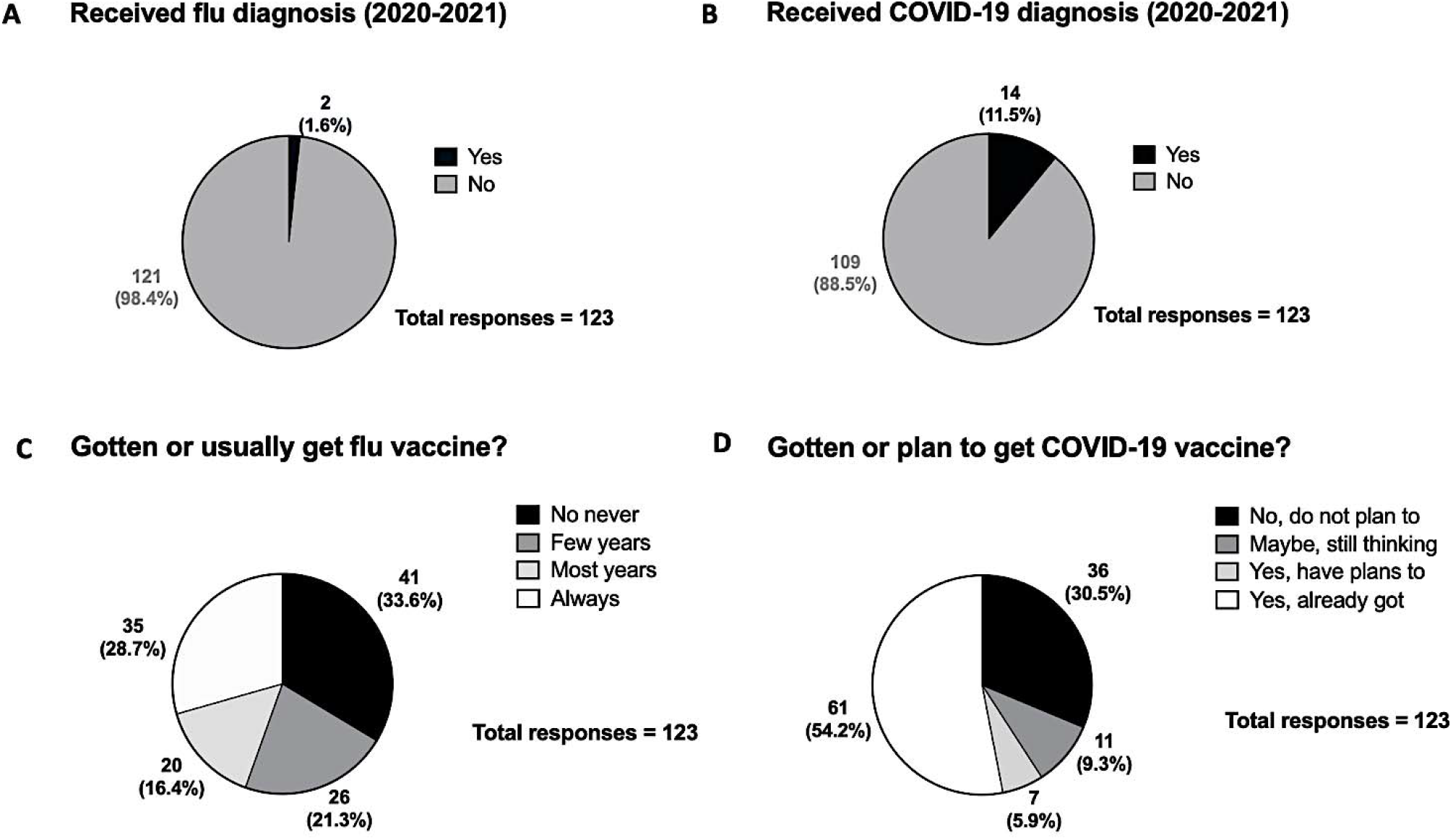
Pie chart representations of frequency of participants having received A) flu diagnosis in the past year, B) COVID-19 diagnosis in the past year, C) flu vaccine, and D) COVID-19 vaccine (or vaccination plans).

We performed numerous bivariate associations to assess demographic factors that were associated with COVID-19 vaccine acceptance (**Table 2**). We found a significant association between COVID-19 vaccine acceptance and age (p<0.01), suggesting that older patients were more likely to be vaccinated against COVID-19 than younger patients were (**Table 2, Figure 3C**). No associations were found between gender, race, or education status and COVID-19 vaccine acceptance (**Table 2**).

**Figure 3.**
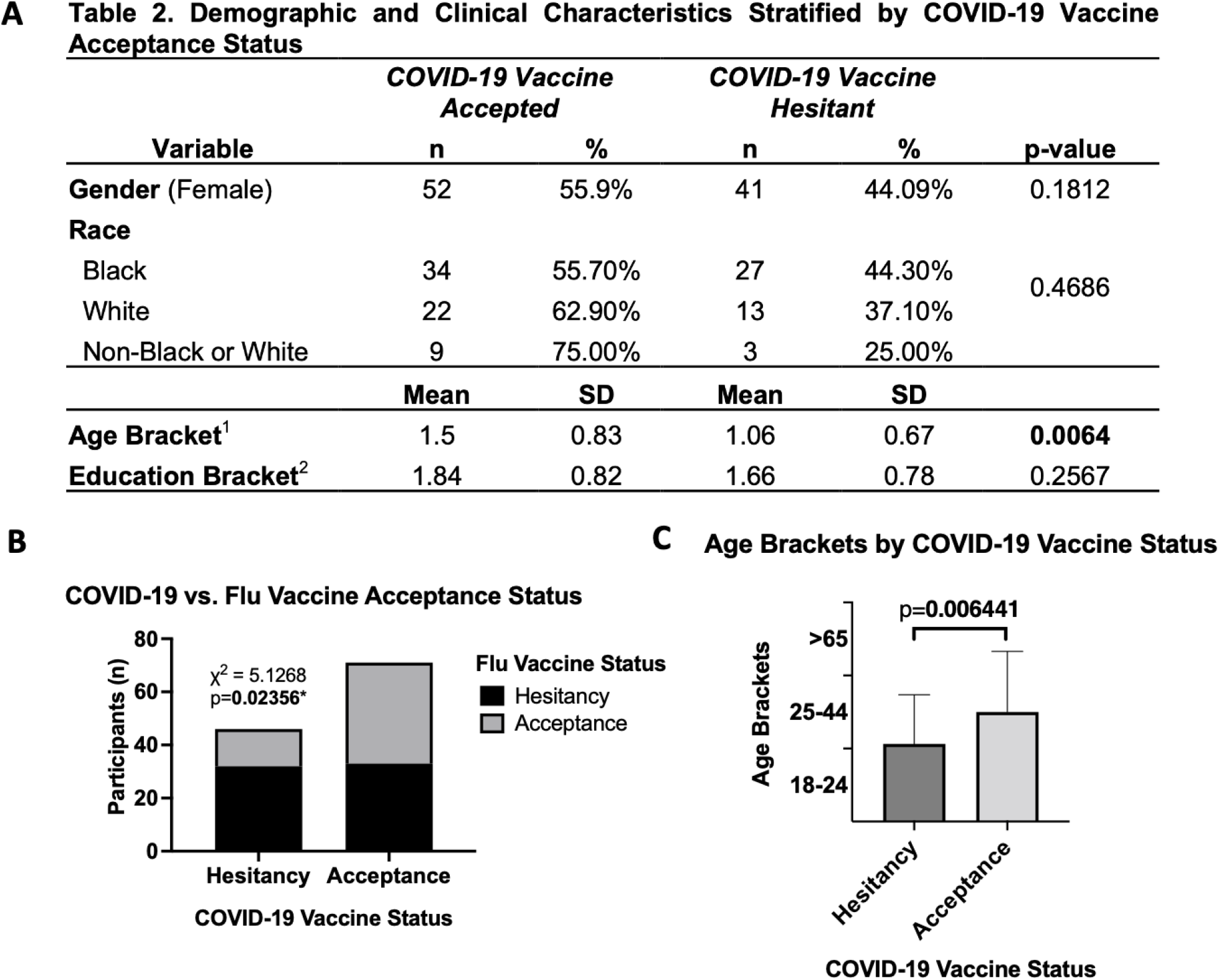
A) Table representation (Table 2) showing bivariate associations of demographic factors with COVID-19 vaccine acceptance. Chi-squared test used for gender analyses, Fisher’s exact test for race analyses, and Mann-Whitney test for age and education analyses. ^1^Age is bracketed by the following code: 0 = 18-24 | 1 = 25-44 | 2 = >65. ^2^Education is bracketed by the following code: 0 = High School - GED | 1 = Associate’s - Some College | 2 = >= Bachelor’s Degree. B) Chi-squared analysis showing association between COVID-19 acceptance vs. flu acceptance statuses. C) Graphical representation of Mann-Whitney test showing significant association between age and COVID-19 vaccine status.

Identical bivariate analyses were run assessing demographic factors versus flu vaccine acceptance. No associations were found (**Table S1, Supplemental**).

We also found that there was a significant association between COVID-19 and flu vaccine acceptance status where, of those who exhibited COVID-19 vaccine acceptance, a significantly higher proportion of them exhibited flu vaccine acceptance compared to the COVID-19 vaccine-hesitant group (χ^2^= 5.1268, p=0.02356) as seen in **Figure 3B**.

In order to understand how participants were staying informed about vaccines and general medical information, we inquired about four domains of information reservoirs. Participants more commonly cited using medical professionals and paper/digital news resources as their primary source for vaccine-specific info compared to word of mouth or social media **(Figure 4A)**. When delineating which of those sources participants trusted the most for medical information, the majority of participants unanimously cited medical professionals as their most trusted source **(Figure 4B)**.

**Figure 4.**
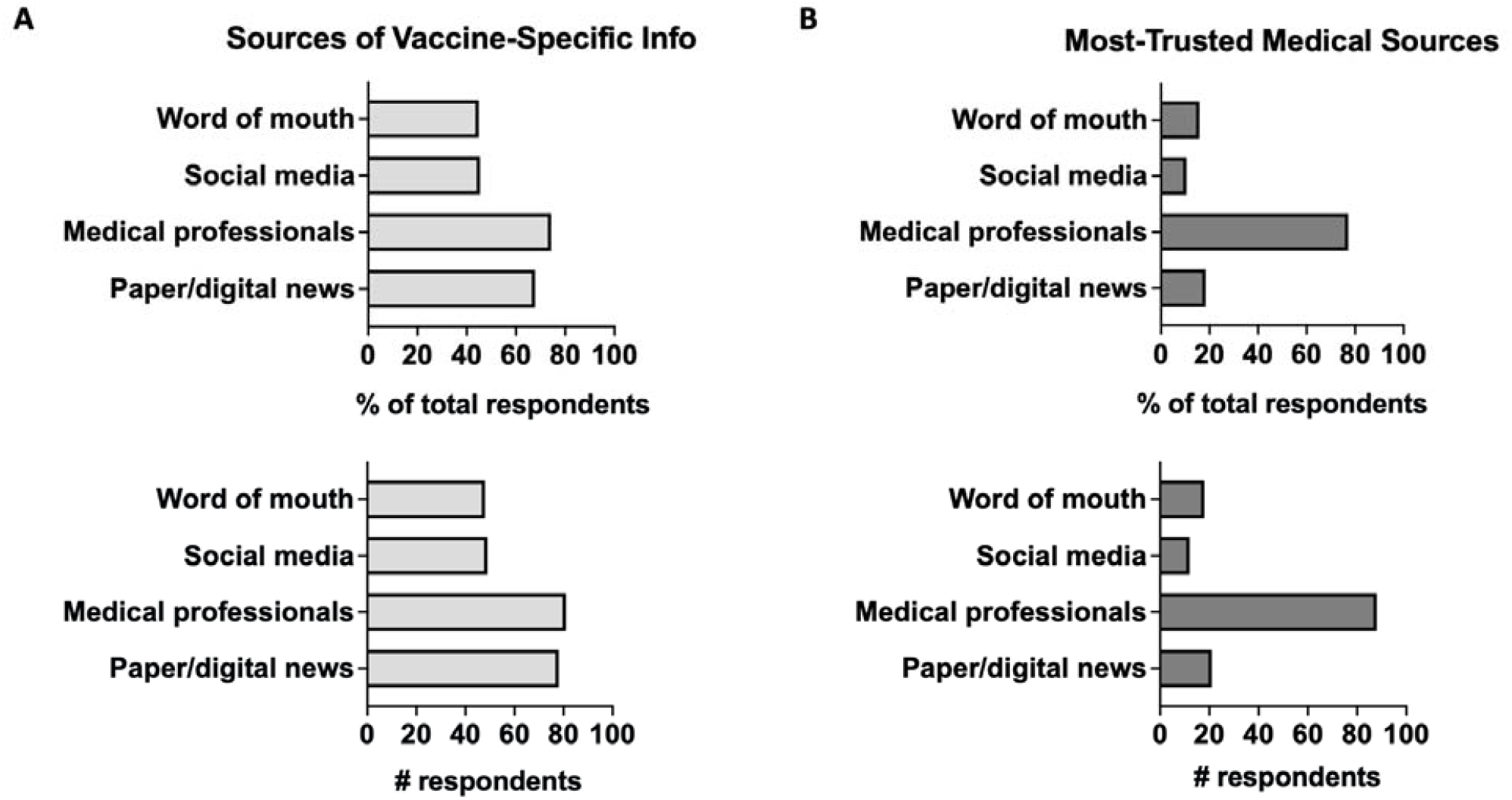
A) Chart representation of trusted sources of vaccine-specific information by frequency (top) and total number (bottom) of respondents per category. B) Chart representation of trusted sources of medical information by frequency (top) and total number (bottom) of respondents. Respondents were prompted to select all source options that applied, and thus may give >1 response.

To assess what vaccine or medical informational sources were associated with higher COVID-19 vaccine acceptance, we ran a multivariable logistic regression. There was no association between the types of trusted sources for vaccine-specific information and actual COVID-19 vaccine uptake **(Table 3A)**. Interestingly, however, when analyzing the sources patients trusted for their overall medical information, all but paper/digital news were significantly associated with COVID-19 vaccine acceptance **(Table 3B)**.

**Table 3.** A) Multiple logistic regression model of trusted vaccine-specific information sources and COVID-19 vaccine acceptance. B) Multiple logistic regression model of trusted medical information sources and COVID-19 vaccine acceptance. Odds Ratio (OR), Confidence Intervals (CI), and p-values reported for each source option.

| **A. Vaccine** |  |  |  |
| --- | --- | --- | --- |
| **Trusted Information Sources** | **OR** | **CI (95%)** | **p-value** |
| Paper or digital news | 0.978 | 0.36 - 2.69 | 0.965 |
| Medical professionals/organizations | 2.064 | 0.78 - 5.64 | 0.148 |
| Social media | 0.438 | 0.17 - 1.11 | 0.085 |
| Word of mouth | 0.518 | 0.19 - 1.42 | 0.200 |
| **B. Medical** |  |  |  |
| **Trusted Information Sources** | **OR** | **CI (95%)** | **p-value** |
| Paper or digital news | 1.352 | 0.37 - 5.97 | 0.665 |
| Medical professionals/organizations | 8.301 | 2.71 - 31.49 | **0.001** |
| Social media | 0.014 | 0.00 - 0.20 | **0.005** |
| Word of mouth | 4.548 | 0.92 - 30.41 | 0.085 |

Trusting medical professionals led to the highest increased odds of getting the COVID-19 vaccine (p<0.001). Word of mouth surprisingly also had a significantly positive relationship with COVID vaccine uptake (p<0.05). Trusting information from social media, on the other hand, was associated with significantly decreased odds of getting the COVID-19 vaccines (p<0.01).

We ran similar multivariable logistic regression to assess what vaccine or medical informational sources were associated with higher flu vaccine acceptance. Medical professionals/organizations was the only trusted source for vaccine-specific information that was significantly associated with flu vaccine uptake (p<0.01) **(Table 4A)**. Trusting medical professionals for general medical information also led to the highest increased odds of getting the flu vaccine (p<0.01) **(Table 4B)**, which was the same trend we saw in COVID-19 vaccination uptake. Unlike with the COVID-19 vaccine, trusting word of mouth or social media for medical information was not significantly associated with flu vaccine uptake.

**Table 4.** A) Multiple logistic regression model of trusted vaccine-specific information sources and influenza vaccine acceptance. B) Multiple logistic regression model of trusted medical information sources and influenza vaccine acceptance. Odds Ratio (OR), Confidence Intervals (CI), and p-values reported for each source option.

| **A. Vaccine** |  |  |  |
| --- | --- | --- | --- |
| **Trusted Information Sources** | **OR** | **CI (95%)** | **p-value** |
| Paper or digital news | 0.535 | 0.20 - 1.45 | 0.216 |
| Medical professionals/organizations | 4.424 | 1.66 - 13.03 | **0.004** |
| Social media | 0.702 | 0.27 - 1.83 | 0.468 |
| Word of mouth | 0.808 | 0.29 - 2.22 | 0.678 |
| **B. Medical** |  |  |  |
| **Trusted Information Sources** | **OR** | **CI (95%)** | **p-value** |
| Paper or digital news | 3.006 | 0.89 - 12.26 | 0.092 |
| Medical professionals/organizations | 5.262 | 1.89 - 17.42 | **0.003** |
| Social media | 0.270 | 0.0317 - 1.69 | 0.190 |
| Word of mouth | 1.076 | 0.26 - 4.41 | 0.916 |

## DISCUSSION

To our knowledge, this study not only represents the first analysis of vaccine uptake in the Turtle Creek community, but one of the first direct assessments of COVID-19 vaccine refusal/hesitancy factors of adults in Western Pennsylvania. Three separate studies of vaccine hesitancy were conducted in nearby Central Pennsylvania, though all reported significant lack of diversity among sociodemographic groups. Study populations were comprised of 92%-97% non-Hispanic white adults, with the majority having attained high income and educational levels (11–13). In our study demographic, majority of participants identified as Black or African American (53%), women (81%), having finished high school and/or some post-graduate work but not college (65%), and in the 25-64 age range (78%), largely representative of known Turtle Creek community demographics (7).

Vaccine uptake information showed that, although this study was conducted prior to the 2021 flu season, only ∼44% of study participants showed predictive flu vaccine acceptance (gets flu vaccine all or most years). This differs from vaccine data gathered in a Central Pennsylvania cohort, which reported a high 88% flu vaccine acceptance rate—highlighting the stark differences that can exist in vaccine uptakes between various Pennsylvania communities and the need for individually assessing smaller local geographical areas (13). Only 54% of participants in the Turtle Creek community had received the COVID-19 vaccine; like in many U.S. communities, this falls short of the national vaccine acceptance rate considered sufficient for herd immunity (described as either >60% or as high as 90% in various studies) (14). As seen with existing U.S. population studies that examined COVID-19 vaccine uptake by demographic association (15,16), our study demonstrates that participants who routinely get flu vaccinations and are older in age were the most likely groups to get vaccinated against COVID-19. This is also in agreement with state-wide data from the PA Department of Health, showing highest vaccination rates among older citizens (>99% vaccinated in 65+ age groups). However, unlike these other studies, our data did not show any associations with gender, race, or education status. While this may be due to our sample size, an important confounder to consider is the health literacy of the Turtle Creek community. Groups most likely to not pursue vaccination are also less likely to be concerned about themselves or family members contracting COVID-19 (1), more likely to believe the virus was manufactured or pandemic exaggerated, hold Republican political beliefs (4,16,17), and have decreased health literacy as demonstrated by knowing less about the safety and science of COVID-19 vaccines (14).

Trusted sources of general medical information and vaccine-specific information are strong predictors of health literacy (11,12,18–20) and were assessed in this study. Medical professionals and paper/digital news were cited to similar frequencies as the top two primary sources of vaccine-specific information for participants; yet, when prompted to share their most-trusted sources of medical information, most participants overwhelmingly cited medical professionals. Medical professionals as a trusted source of both vaccine-specific and overall medical information were positively associated with flu vaccine acceptance. However, with regards to COVID-19 vaccine uptake and trusted sources for information, it appears that people’s *overall* belief in medical professionals vs. other news sources influenced their vaccine choice, but their trust in those sources’ ability to provide info about *vaccines specifically* did not correlate with higher or lower vaccine acceptance rates. Several factors may lead to a loss of correlation between vaccine acceptance and trusting news sources for vaccine-specific information, including what a participant’s definition of vaccine-specific information may be (i.e. any information related to vaccines versus research articles specifically), lack of access to health centers or vaccine-related health information, lack of public accessibility for vaccine-specific research data, and/or less willingness or motivation to seek vaccine-specific resources.

Interestingly, when examining the correlation between most trusted sources of overall medical information and vaccine acceptance, all news sources except paper/digital news were significantly associated with COVID-19 vaccine uptake. This differed from flu vaccine uptake, which was associated with paper/digital news and medical professionals but not social media or word of mouth. Yet, news sources are still an important outlet on which to improve vaccine information, as 43% and 45% of U.S. adults report frequently receiving their news online or on a mobile device, respectively (21). The correlation between trusted sources of medical information and vaccine acceptance also revealed trusting information from medical professionals was most strongly associated with both COVID-19 and flu vaccine uptake. This corroborates findings from other studies which list doctors and medical professionals among the most trusted sources of medical information during the COVID-19 pandemic (18,19). Of note, lack of trust in medical professionals is significantly associated with lower health literacy, while routine use of medical websites is associated with higher literacy. Thus, it is critical to improve trust in medical professionals and access to medical information in vaccine-hesitant populations. Word of mouth from friends and family was also positively associated with COVID-19 vaccine uptake, although few studies of COVID-19 news sources have included this variable. Literature on the association between word of mouth/friends and health literacy is mixed, with some studies reporting positive correlations to vaccine literacy and others reporting negative correlations (18,19). This may be, in part, linked to the heterogeneity of a person’s friend group and whether their perspectives differ or correlate to the participant’s own views on vaccines. More studies will be needed to accurately define relationships between COVID-19 vaccine acceptance and friends as a source of information.

Trust in social media for medical information was associated with decreased likelihood of COVID-19 vaccine acceptance. This is supported by existing studies, which show that use of social media for health information was inversely associated with flu vaccine uptake (22) and correlated to lower health literacy (19). However, this data should not dismiss social media as a highly relevant source for disseminating accurate vaccine information. Exposure to vaccine-critical/anti-vaccination websites directly decrease intentions to vaccinate (23–25). Social media is a frequently utilized news source, and, as aptly described by Wolynn & Hermann, “When health care providers and vaccine advocates go silent on social media, they create a vacuum—one that antivaccine voices are only too happy to fill” (26). With the prevalence of misinformation, it is important to address patient’s concerns and questions to promote the uptake of these vaccines and to combat the spread of COVID-19. A study from 2018 found that attitudes towards vaccines were more positive when social media posts are associated with fact-checking labels and trusted institutions/organizations. The study highlighted the importance of addressing misinformation in social media and the potential to collaborate between social media platforms and public health entities to spread factual information (27).

In conclusion, our study uncovered significant trends in flu and COVID-19 vaccine uptake, sociodemographic associations with vaccine acceptance, most used and trusted medical news sources, and reasons for flu and COVID-19 vaccine resistance/hesitancy in the Turtle Creek community. A significant strength of our study is that, unlike other surveys of nearby Pennsylvania counties, our study population includes substantial racial diversity, economic diversity, and urban representation. Another strength is that, while much of the research on COVID-19 vaccine research was predictive in nature (being conducted when COVID-19 vaccines were not yet developed/available to the public), our study was conducted at a time of widespread accessibility to COVID-19 vaccinations. Limitations included a small sample size, the inability to directly assess 2021 flu season vaccination rates in our time frame, and the focused study of adult patients of an ambulatory practice (excluding pediatric populations and adult patients who receive healthcare in facilities from our assessment). Our study’s cross-sectional nature also contains the inherent limitation of demonstrating only disease association and not causation. Importantly, our study elucidated clear differences in COVID-19 and flu vaccination trends and sociodemographic makeup between the Turtle Creek clinic and existing studies conducted in nearby PA counties, PA state data, or U.S. population studies, highlighting the inability to base vaccine policies on generalized national assessments and the importance of collecting more geographically detailed vaccine data that includes information on diverse populations.

## SUPPLEMENTAL FIGURES

**Table S1.** Demographic and Clinical Characteristics Stratified by Flu Vaccine Acceptance Status. A) Table representation (Table 2) showing bivariate associations of demographic factors with influenza vaccine acceptance. Chi-squared test used for gender and race analyses, and Mann-Whitney test for age and education analyses. ^1^Age is bracketed by the following code: 0 = 18-24 | 1 = 25-44 | 2 = >65. ^2^Education is bracketed by the following code: 0 = High School - GED | 1 = Associate’s - Some College | 2 = >= Bachelor’s Degree.

| **Variable** | ***Flu Vaccine Accepted*** |  | ***Flu Vaccine Hesitant*** |  | **p-value** |
| --- | --- | --- | --- | --- | --- |
|  | **n** | **%** | **n** | **%** |  |
| **Gender (Female)** | 46 | 47.9% | 50 | 52.08% | 0.5987 |
| **Race** |  |  |  |  |  |
| Black | 26 | 41.93% | 36 | 58.07% | 0.2774 |
| White | 18 | 51.43% | 17 | 48.57% |  |
| Non-Black or White | 9 | 35.71% | 5 | 64.29% |  |
|  | **Mean** | **SD** | **Mean** | **SD** |  |
| **Age Bracket<sup>1</sup>** | 1.364 | 0.85 | 1.29 | 0.76 | 0.5982 |
| **Education Bracket<sup>2</sup>** | 1.736 | 2.66 | 1.778 | 2.34 | 0.6775 |

### APPENDIX

#### A.1 Full Phone Survey Questionnaire

1. Are you okay with taking a few minutes to talk with me today to discuss how you feel with vaccines and about your history with vaccines?
  a. Yes
  b. Maybe
  c. No
2. Have you been diagnosed by a medical professional with the flu in the fall 2020-until now?
  a. Yes
  b. Maybe
  c. No
3. Have you ever tested positive for COVID19 and/or been diagnosed by a medical professional in fall 2020-until now?
  a. Yes
  b. Maybe
  c. No
4. Do you usually get a flu shot?
  a. Always
  b. Most years
  c. Few years
  d. No never
  e. I don’t want to answer this question
5. What is your reason for the question above?
  a. Example: Why do you always get it?
  b. Example: Why do you get it sometimes but not other times?
  c. Example: Why do you not get it? Are there specific barriers you face?
6. Have you gotten the COVID-19 vaccine or have plans to get it?
  a. Yes, I already got it
  b. Yes, I have plans to get it
  c. Maybe, I’m still thinking about it
  d. No, I don’t have plans to get it and I don’t want it
  e. I don’t want to answer this question
7. What is your reason for the question above?
  a. Example: Why did you get it? / Why do you want to get it?
  b. Example: Why are you hesitant about getting it?
  c. Example: Why do you not want to get it? Are there specific barriers you face? What are you worried most about?
8. From where do you receive information about vaccines?
  a. Paper or digital news
  b. Social media from non-governmental and non-medical/scientific accounts (ex. friends, bloggers, celebrities, etc.)
  c. Word of mouth from non-medical people
  d. Medical professionals/health organizations in person and/or on social media
  e. Other
9. What sources of information do you trust the most for medical information, in general (i.e. not specific to vaccines)?
  a. Paper or digital news
  b. Social media from non-governmental and non-medical/scientific accounts (ex. friends, bloggers, celebrities, etc.)
  c. Word of mouth from non-medical people
  d. Medical professionals/health organizations in person and/or on social media
  e. Other
10. Patient age
  a. 18-24 years
  b. 25-44 years
  c. 45-64 years
  d. 65 years and over
11. What gender identity do you most identify with?
  a. Female
  b. Male
  c. Transgender Female
  d. Transgender Male
  e. Gender-variant/non-conforming
  f. Another option. not listed here (please specify)
  g. Prefer not to answer
12. What race/ethnicity do you most identify with?
  a. American Indian or Alaska Native
  b. Asian or Asian American
  c. Black or African American
  d. Hispanic, Latino, Latina, or Latinx
  e. Middle Eastern or Northern African
  f. Native Hawaiian or Other Pacific Islander
  g. White
  h. Another option not listed here (please specify)
  i. I prefer not to answer this question
13. What is the highest degree or level of education you have *completed*?
  a. Some high school
  b. High school
  c. GED
  d. Associate’s degree
  e. Bachelor’s degree
  f. Master’s degree
  g. Professional or doctoral degree or higher
  h. Trade school
  i. Other (please specify)
  j. Prefer not to say

